# Wearable Sensor and Digital Twin Technology for the Development of a Personalized Digital Biomarker of Vaccine-Induced Inflammation

**DOI:** 10.1101/2024.01.28.24301887

**Authors:** Steven R. Steinhubl, Jadranka Sekaric, Maged Gendy, Huaijian Guo, Matthew P. Ward, Craig J. Goergen, Jennifer L. Anderson, Sarwat Amin, Damen Wilson, Eustache Paramithiotis, Stephan Wegerich

## Abstract

Effective response to vaccination requires activation of the innate immune system, triggering the synthesis of inflammatory cytokines. The subjective symptoms related to this, referred to as reactogenicity, affect a variable percentage of vaccinated people to different degrees, with evidence supporting a relationship between the severity of symptoms a person experiences and their eventual immune response. Wearable sensors allow for the identification of objective evidence of physiologic changes a person experiences in response to vaccine-induced inflammation, but as these changes are subtle, they can only be detected when an individual’s pre-vaccination normal variability is considered. We used a wearable torso sensor patch and a machine learning method of similarity-based modeling (SBM), which learns the dynamic interplay between multivariate input sources, to create a physiologic digital twin for 88 people receiving 104 vaccine doses. By effectively removing expected variations and leaving only vaccine-induced differences, we developed a multivariate digital biomarker that incorporates changes in multiple continuously monitored physiologic data streams to measure the degree and duration of vaccine induced inflammation. This objective measure correlated with subjective symptoms, and in a 20-person subset, both humoral and cellular immunogenicity.

## Introduction

Immune cell activation is essential to a successful vaccination strategy, with inflammation being the immune system’s initial response to this activation. The level of vaccine-induced inflammation plays an essential role in its tolerability and potentially its efficacy.^1,2^ Presently, the symptomatic tracking of reactogenicity – the physical manifestations of vaccine-related inflammation – is the only measure of inflammation that is monitored at any scale.^3^ For example, following the second dose of a mRNA based COVID-19 vaccine nearly 70% of individuals participating in the Centers for Disease Control and Prevention’s (CDC) v-safe study (<5% of all vaccinated individuals in the US) reported having a systemic symptom such as fatigue, myalgias or chills.^4^ However, the subjective nature of these data limit their value as a measure of individual inflammation as they are susceptible to a nocebo effect. An analysis of the placebo arms of COVID-19 vaccine randomized trials found that 76% of systemic symptoms experienced after the first dose, and 52% after the second dose could be attributed to the nocebo effect.^5^

Currently, objective measures of vaccine-induced inflammation are dependent on intermittent and infrequent sampling of blood-based soluble factors such as chemokines and cytokines.^6^ Most, but not all, of these studies have found significant relationships between blood-based inflammatory biomarkers and measures of reactogenicity and/or immunogenicity following vaccination against COVID-19 or other pathogens.^1,2,7–9^ However, due to the invasive requirements of these studies, sample sizes are limited, and the frequency and duration of testing is minimized. Non-invasive sources of soluble inflammatory biomarkers that would allow for the capture of the fuller extent of an individual’s inflammatory response to a vaccine, such as urine and saliva, have been evaluated, but have yet to be proven effective.^6,10^

A potentially novel approach to quantifying the totality of an individual’s inflammatory response to vaccination could be through wearable sensors that can continuously track individual physiologic and behavioral changes following vaccination to create a digital biomarker.^11^ Recently, a range of wearable sensors – wrist wearables, rings and torso patches - have been shown to be able to detect the subtle physiologic changes associated with COVID-19 vaccination-induced inflammation.^12–17^ The degree of changes are so small that without knowledge of a person’s unique pre-vaccine normal levels and natural variability, the detection of subtle deviations associated with vaccine-induced inflammation would not be possible. Changes that, for example, might be only two beats per minute difference in resting heart rate – a change that would never be detectable on a population level due to marked inter-individual variability in resting heart rate.^12,18^ As much of this prior work has utilized consumer devices, physiologic and behavioral changes following vaccination have mostly been determined based on a single daily summary value for each parameter, with most physiologic measures determined during sleep. Even with this limited data density, the one study that explored immunogenicity did find a significant correlation between several physiologic measures and semi-quantitative antibody levels.^13^

In the present study, we sought to develop a personalized digital biomarker for COVID-19 vaccine-induced inflammation utilizing several advanced technologies; a medical grade patch biosensor able to continuously capture multiple parameters, and an analytics platform using a machine learning method of similarity-based modeling (SBM), which learns the dynamic interplay between multivariate input sources.^19^ Combined, these technologies enabled the development of personalized pre-vaccine baseline models of each participant’s unique physiologic dynamics – a physiologic “digital twin.”^20^ When these models were then applied to post-vaccination data to remove expected individual variations, continuous vaccine-induced changes were able to be isolated to create a multivariate, personalized biomarker of inflammation – an inflammatory multivariate change index (iMCI).

Here we report the measured inflammatory response using this new iMCI digital biomarker in 88 individuals undergoing voluntary vaccination for COVID-19 in a real world setting for a total of 104 doses. We quantify the relationship between that measure and subjective symptoms in all individuals, finding a statistically significant correlation. In addition, in a subset of participants, we identified a statistically significant relationship between iMCI and the humoral and cellular immune responses early following vaccination. These results provide preliminary support for the potential value of a personalized digital biomarker to identify individual inflammatory response during immune system activation.

## Methods

### Clinical Setting

Participants were recruited through one of two study protocols. The majority of participants were enrolled through the Vaccine Induced Inflammation Investigation (VIII) study protocol. The second protocol was the Continuous Physiologic Monitoring for Immune Response via Wearable Sensor Data (COmMON SENS) study. The VIII study was approved by the Sterling IRB (ID# 8842-SRSteinhubl) in April of 2021. Clinicaltrials.gov registration number was NCT05237024. The COmMON SENS study was approved by the Purdue University IRB (ID# IRB-2021-453) April, 2021.

For the VIII study, individuals who were already voluntarily planning to receive a vaccine against COVID-19 were recruited from the general population primarily via email outreach disseminated by employees of the study sponsor, physIQ (Chicago, IL, USA, but since purchased by Prolaio, Scottsdale, AZ, USA). Employees were encouraged to further disseminate recruitment information to their family and friends. The choice by any potential participant to get a vaccine was entirely voluntary and only people already planning to be vaccinated were approached to enroll in the study. Participants in the immunogenicity substudy of the VIII study were recruited in a similar manner with outreach disseminated by CellCarta (Montreal, Qc, CA), the cellular immunogenicity lab.

In the COmMON SENS study, Purdue University students, staff, and faculty were recruited via advertising to the general university population, affiliates, and friends. Again, only individuals who were voluntarily planning to receive a COVID-19 vaccine were recruited.

### Inclusion & Exclusion Criteria

Any individual over age 18 (participants between age 12 and 17 were allowed in the VIII study, although none were recruited) who was planning to receive any of the 3 FDA-approved vaccines against COVID-19 were eligible for enrollment. The only exclusion criterion was known allergy to the adhesive of the sensor patch.

### Study Methods

Individuals enrolled were asked to wear a patch sensor for ∼12 to14 days surrounding vaccination. Participants could agree to monitor themselves during more than one vaccine dose. Volunteers were asked to place the patch on themselves and begin monitoring 5 days prior to their planned vaccination and continuing for a total of up to 14 days. As the battery life of each disposable patch was ∼7 days, each participant was asked to sequentially wear two patches at the time of each vaccine.

All participants received a locked-down Android phone with a preloaded app to enable patch and survey data capture. The app enabled participants to mark the day and time they received each vaccine dose and respond to daily survey questions for up to 7 days following vaccination to track all subjective symptoms.

### Immunogenicity Sub-Study

Participants in the immunogenicity sub-study followed the identical protocol as all study participants with the exception of agreeing to a series of blood draws. For participants who had yet to receive their first vaccination (n=3), there were 4 scheduled blood draws. The first ∼5 days prior to their first vaccine, the second ∼14 days after their first dose, the third ∼14 days after their second dose, and their final blood draw ∼60 days after their second vaccine dose. The majority of participants underwent only 3 blood tests -∼5 days before their second or third vaccine dose, and again ∼14 days and ∼60 days after that dose.

### Wearable Sensor

The VitalPatch^TM^ by VitalConnect (San Jose, CA) is an FDA 510(k)-cleared, wearable, disposable adhesive patch with an integral one-time use battery and integrated electronics. The battery life of each patch lasts 7 days. The patch was self-applied by the participant to their left upper chest. Guidance for placement and confirmation of connectivity was provided via the app.

The patch transfers biosignal data over Bluetooth low energy protocol to the mobile app. Once the mobile app has received the biosignals from the VitalPatch^TM^, the biosignals are uploaded to physIQ’s cloud-based server using digital cellular or Wi-Fi networks. No personal identifiers were stored or transmitted with the data either from the sensor or from the mobile app. Upload via digital cellular network was secured with Transport Layer Security (TLS) cryptographic protocol between the mobile phones and the server. The physIQ platform was securely hosted in the Google cloud and the analytics server stored the raw physiological telemetry data captured by the study device. All the telemetry waveform data were stored only by participant ID. The data could only be obtained or viewed via secure authenticated login.

### physIQ Platform and Personalized Physiology Analytics

The cloud-based analytics platform used a general machine learning method of similarity-based modeling (SBM), to analyze collected data. SBM models the behavior of complex systems (e.g., aircraft engines, computer networks, or human physiology) by learning tandem patterns among system variables as they are periodically sampled together.^19^ Personalized baseline models of each participant’s unique physiological dynamics are established, creating a “digital twin,” which when compared to new input data following a possible immune-stimulator, removes expected variations, leaving only inflammation-induced differences. These are the residuals. These residuals are combined into an inflammation Multivariate Change Index (iMCI), which is updated on a 15-minute basis, allowing for the tracking of the onset, offset and degree of inflammatory change. Patch data was filtered for quality using an ECG-based signal quality index (SQI), that is graded 0-1 with 1 as highest quality.

#### Uni-parametric Analysis

Patch ECG-derived cardiorespiratory features were filtered with a threshold of SQI > 0.9. Since activity level and skin temperature derived from the patch are not related to the ECG signal, they were not filtered with SQI, however any skin temperature values below 33 C and above 42 C were ignored. Five, 3-hour aggregate parameters were derived from 1-minute derived features after baseline normalizing using z-scoring on an individual participant basis. The mean and standard deviations used for the z-scoring, were calculated from feature data generated during the 48-hour window prior to a vaccine dose. The corresponding post vaccine dose aggregate data for each participant were used to identify cases where at least one standard deviation of change from baseline occurred. .

#### Multiparametric (iMCI) Analyses

The multivariate iMCI is a personalized modeling algorithm that uses coincident heart rate, heart rate variability, respiration rate, activity level and skin temperature derived from the patch device as input. A unique model was trained for every participant based on data collected prior to vaccine doses. For data to be used as input to iMCI, activity level had to be less than 0.05 g (a level corresponding to normal walking), skin temperature between 30° C and 40° C, and SQI ≥ 0.9. Additionally, during training, heart rate was required to be between 40 bpm and 250 bpm, and respiration rate between 8 and 35 breaths per minute. To train an individual iMCI model, it was required that 2500 1-minute samples of input variables be available prior to vaccine doses and that 1-minute samples prior to vaccine were distributed over 3 days.

#### iMCI Total Response

The metric defined for assessing iMCI total inflammatory response (“iMCI Total Response”), employed an area under the curve (AUC) approach. The metric is defined for a fixed window of time (Y) starting from the time of administration of a vaccine dose. The time window used was Y = 72 hours. The iMCI Total Response was defined as A_i_/A_T_ as illustrated in **Supplemental Figure A**, where A_T_ is the total rectangular area within the time window and A_i_ is the area under the curve for iMCI during the window.

#### iMCI Detectable Response

A detectable response was defined as a collection of iMCIs with persistent, non-zero iMCI values within the fixed 72-hour window following a vaccine. The 72-hour window was selected to capture as much of the vaccine-induced inflammatory response and minimize any potential noise. In addition to being a persistent trend (non-zero iMCI trend > 1 hour), the iMCI response had to satisfy conditions that were designed to rule-out small magnitude, random fluctuating trends that are likely due to noisy arbitrary inputs. Detectable response was defined as the presence of at least 50% of 15-minute steps within a 6-hour sliding window with an iMCIs > 0.10. If any of such occurrences has been found in the fixed 72-hour window following a vaccine the individual has a detectable inflammatory response. In this way we identified 66 (63.5%) vaccine doses for which an inflammatory response was of detectable size. To estimate the false positive rate of an algorithm that identifies a detectable response we select an independent data consisting of N_C_ = 76 participants from two different, healthy, non-vaccine, non-infection cohorts to serve as a control group. These data were processed in the same way as the current data set to generate iMCIs. From each of N_C_ participants we randomly selected a W_C_ = 100 of 72-hour chunks of iMCI data and tested for a presence of a 6-hour sliding window (in steps of 15-minutes) with at least 50% of iMCIs > 0.10. Each chunk of 72-hour iMCI data was labeled as a positive or negative decision depending on whether such a window has been found or not. In each of 1000 bootstraps we pull N_C_ x W_C_ estimated decisions (one per each vaccine dose) and compare them to decisions labeled as 76 negative for control group and 66 positive from the current study cohort to estimate the performance of “detectable response”-identification algorithm and corresponding 95% confidence intervals (CI); True Positive Rate (TPR) = 100%, Specificity (SPC) = 67.7% (67.4%, 68.0%), False Positive Rate (FPR) = 32.3% (32.0%, 32.6%), Positive Predictive Value (PPV) = 73.0% (72.8%, 73.2%), Negative Predictive Value (NPV) = 100%, Acceptance (ACC) = 82.7% (82.5%, 82.9%).

### Immunogenicity Studies

Initial processing of blood samples to isolate peripheral blood mononuclear cells (PBMCs) and blood plasma occurred within 2 hours of blood draw. When possible, 1.5 ml of plasma was removed from the top of each spun sample tube prior to buffy coat isolation. Aliquots of plasma were stored at -80°C. PBMCs, with a target concentration of 10.0x10^6^ PBMC/ml/vial, were cryopreserved in liquid nitrogen until batch analysis.

#### Flow cytometry-based T-cell assays

Intracellular cytokine stain assay was performed at CellCarta Bioscience, Inc. (Montreal, QC, Canada) similar to as previously described.^21^ For each sample, 4 conditions were used: DMSO, S peptide small pool, non-S peptide pool and staphylococcal enterotoxin B as a positive control. PBMCs were rested and then stimulated for 16–18 h at 37 °C, 5%CO2 in the presence of secretion inhibitors. After the stimulation, cells were stained with fixable Aqua dead cell stain as well as surface antibodies, followed by intracellular staining with cytokine (e.g. IFNγ, IL-2, IL-4 and IL-21) or cytotoxicity marker (perforin) using BD Cytofix/ Cytoperm protocol. Samples were acquired on a BD Fortessa X20 cytometer and data was analyzed using CellEngine software. The frequency of cytokine-producing antigen specific T-cells was determined by subtraction of the background cytokine response in unstimulated control samples from the positive response in the samples stimulated with SARS-CoV-2 peptide pools. All negative values after subtraction of background were set to 0.

#### Anti-spike IgG

Humoral immunogenicity was determined from plasma samples and defined as SARS-CoV-2 anti-spike IgG titers. Anti-spike IgG concentrations were determined by ELISA (reported as ELISA laboratory units [ELU]/mL) at Nexelis (Laval, QC, Canada) similar to as previously described.^22^

### Statistical Analyses

#### Population level daily summary changes

To test for a difference in the pre- and post-vaccination levels for physiologic and behavioral biometrics over the entire study population, medians were calculated using all available pre-vaccine data and 5-day post vaccine data.

Statistically significant differences were determined based on Wilcoxon signed rank test.

#### Post-vaccine inflammatory response by vaccine and individual characteristics

The probabilities that the two data sets come from different continuous distribution at the 5% significance level are obtained using Kolmogorov–Smirnov (KS) test.

#### Relationship of iMCI to Subjective Reactogenicity

We compared the AUC iMCI between the two populations – those that reported systemic symptoms and those that reported have no symptoms or local symptoms only - using a two-sample Kolmogorov-Smirnov goodness-of-fit hypothesis test (KS test) to see if there is a significant difference between the two populations in inflammatory response.

#### Relationship of iMCI to Immunogenicity

The baseline-subtracted amounts of anti-spike protein IgG and T-cell assays CD4+/IL-21+ and CD8+/IFNγ+ obtained from blood samples drawn at day 14 after vaccine was administered were compared to AUC iMCI to assess the Spearman correlation between the two. In case of 2^nd^ doses, the baseline values are those obtained prior to vaccine dose two (post 1^st^ vaccine) while in case of 3^rd^ doses the baseline values were obtained from blood samples drawn prior to third dose (post 2^nd^ vaccine).

One participant was excluded due to insufficient wearable data. The reasoning for removal of another 3 participants’ data in evaluating the relationship between AUC iMCI and cellular immunogenicity and 2 participants’ data for humoral immunity is explained in **Supplemental Figure B.**

The fitted lines were obtained with a robust fit method which is an alternative regression in the presence of outliers or influential observations making it less sensitive to outliers than standard linear regression. In cases (a) and (b) of Figure 5 we used Welsch weight function and for (c) we used Andrews weight function, all with the same tune parameter 0.8. The choice of tuning parameter affects the number of data points used in fitting procedure and the model statistics. Each data point is weighted differently based on how it affects the model statistics i.e., based on the magnitude of the residual for that data point.

## Results

### Participants

A total of 107 individuals consented to participate and included 137 vaccine doses. After excluding individuals with insufficient baseline or post-vaccine data, 88 participants were included in this analysis with a total of 104 vaccine doses. Forty-two (47.7%) were female and the mean age (±SD) for analyzed population was 37.9 (±13.9) years with a range of 19 to 69 years. Eleven people (10.6%) self-reported prior COVID infection. All participants but one received one of the two available mRNA COVID-19 vaccines – Moderna’s mRNA-1273 (43 doses), Pfizer-BioNTech’s BNT162b2 (48 doses) and for 12 doses participants were not sure which mRNA vaccine they received. One person received the Janssen viral vector vaccine.

Response to the first vaccine dose (including the 1 Janssen vaccine recipient) was monitored in 15 participants, to the second dose in 44 individuals, and a third dose in 45 people.

Fourteen people provided data around 2 different vaccine doses, and one person for three doses.

### Wearable Data

The torso ECG patch was worn for a mean (±SD) of 4.2 (± 2.1) days prior to vaccination and 7.6 (± 3.0) days after. A total of 37,279 hours of data were analyzed out of a possible 38,832 hours of total patch wear time (96% data availability).

At a population level, small but significant changes relative to individual pre-vaccine baselines, consistent with a physiologic response to inflammation, were detectable in heart rate (HR), skin temperature, heart rate variability (HRV) and respiratory rate (RR) for up to 3 days following vaccination. **(Figure 1)**

**Figure 1.**
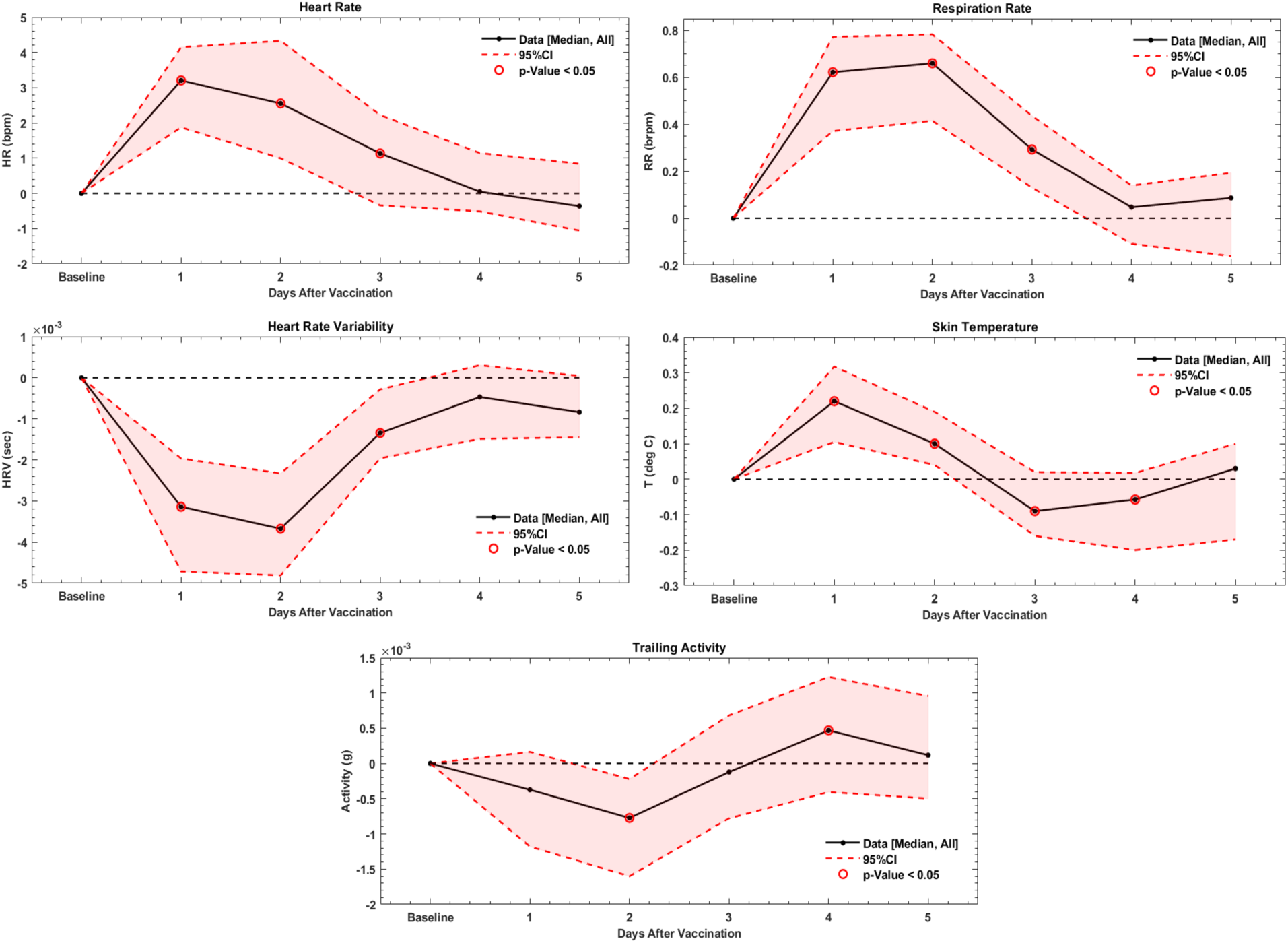
Population level daily summary changes in heart rate (HR), heart rate variability (HRV), respiratory rate (RR), activity level (labeled as “trailing activity”, which indicates that the activity level is filtered using a 20-second moving average at the 1-minute level to remove higher frequency variability over time), and skin temperature following any vaccination dose relative to pre-vaccine baseline with significant changes circled in red. HR changes are in beats per minute, HRV in seconds, RR in breaths per minute and skin temperature in ^o^C.

### Uni-parametric Physiologic Changes from Baseline

Individual differences in changes in single physiologic parameters following vaccination were evaluated by tracking deviations in each z-scored measured parameter relative to each participant’s 48-hour pre-vaccine dose baseline period. Among the 85 participants who had data surrounding a second or booster dose, detectable changes greater than one standard deviation relative to an individual’s baseline were seen in 28 (33%) of individuals in skin temperature, 24 (28%) in HR, 11 (13%) in RR, 7 (8%) in HRV, and 1 (1%) in activity level. In total, a change in one or more individual parameters of one standard deviation or greater was detected in 46 (54%) of participants following a second or booster vaccine dose.

As shown in **Figure 2**, there was significant variability in the onset, duration, and degree of change between individuals in each parameter. In addition, a person’s response in one parameter did not necessarily predict their response in another parameter. For example, participant 024 experienced relatively large changes in all parameters except for activity level, whereas participant 004 experienced a large change in only respiratory rate with a moderate change in skin temperature. Also, participant 023 experienced a large change in HRV and skin temperature, but only low to moderate change in HR.

**Figure 2.**
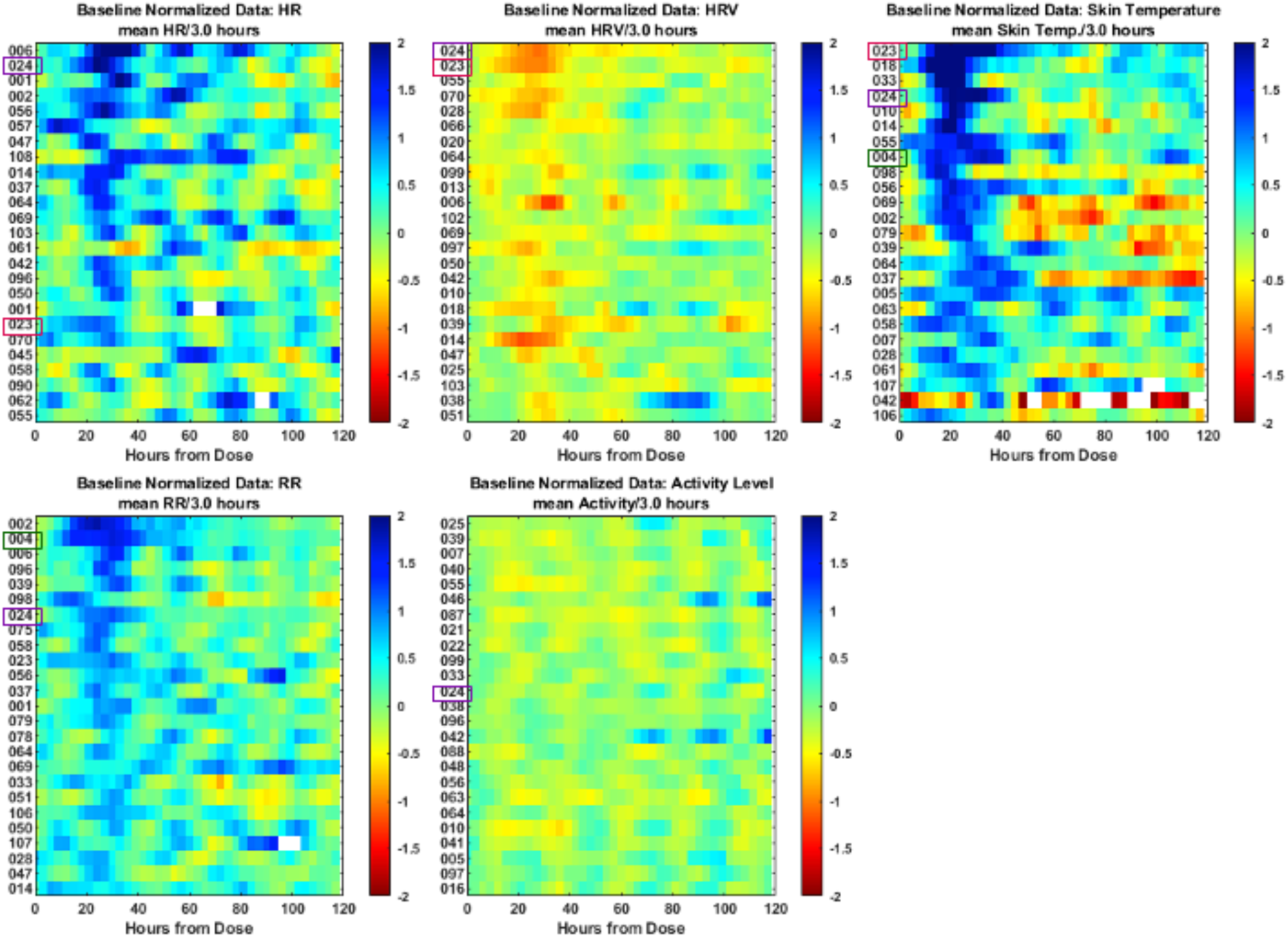
Heat maps of the 25 participants with the greatest changes relative to their pre-vaccination baselines in each of 5 individual parameters in order of degree of change from higher to lower. Several participants (numbers 04, 23 and 24) are highlighted as examples of how the relative change in one parameter is not predictive of the change in another.

### Inflammatory Multivariate Change Index

The individual differences in multiple parameters are combined into an inflammatory Multivariate Change Index (iMCI), which is a personalized modeling algorithm that uses coincident heart rate, heart rate variability, respiration rate, activity level and skin temperature derived from the patch device as input and is further detailed in the Methods section. To best quantify an individual’s total vaccine-associated inflammatory response, the area under the iMCI curve (AUC iMCI), which encompasses the duration and degree of measured physiologic changes following vaccination, was determined for each participant’s post-vaccine experience beginning immediately after vaccination up to 3 days (72 hours) after. **(Supplementary Figure A)**

The first vaccine dose was associated with a less pronounced AUC iMCI response relative to those receiving their 2^nd^ or 3^rd^ dose. **(Table 1 and Figure 3a)** Sixty-five percent of second or booster doses led to a detectable increase in iMCI after vaccination compared to only 53% of those after a first dose, with ‘detectable’ as defined in the Table and in the Methods section. The total response in participants who received the mRNA-1273 vaccine tended to be greater and appeared to be of longer duration **(Figure 3b)** as measured by continuous iMCI than those treated with BNT162b2, although between group differences were not significant.

**Figure 3.**
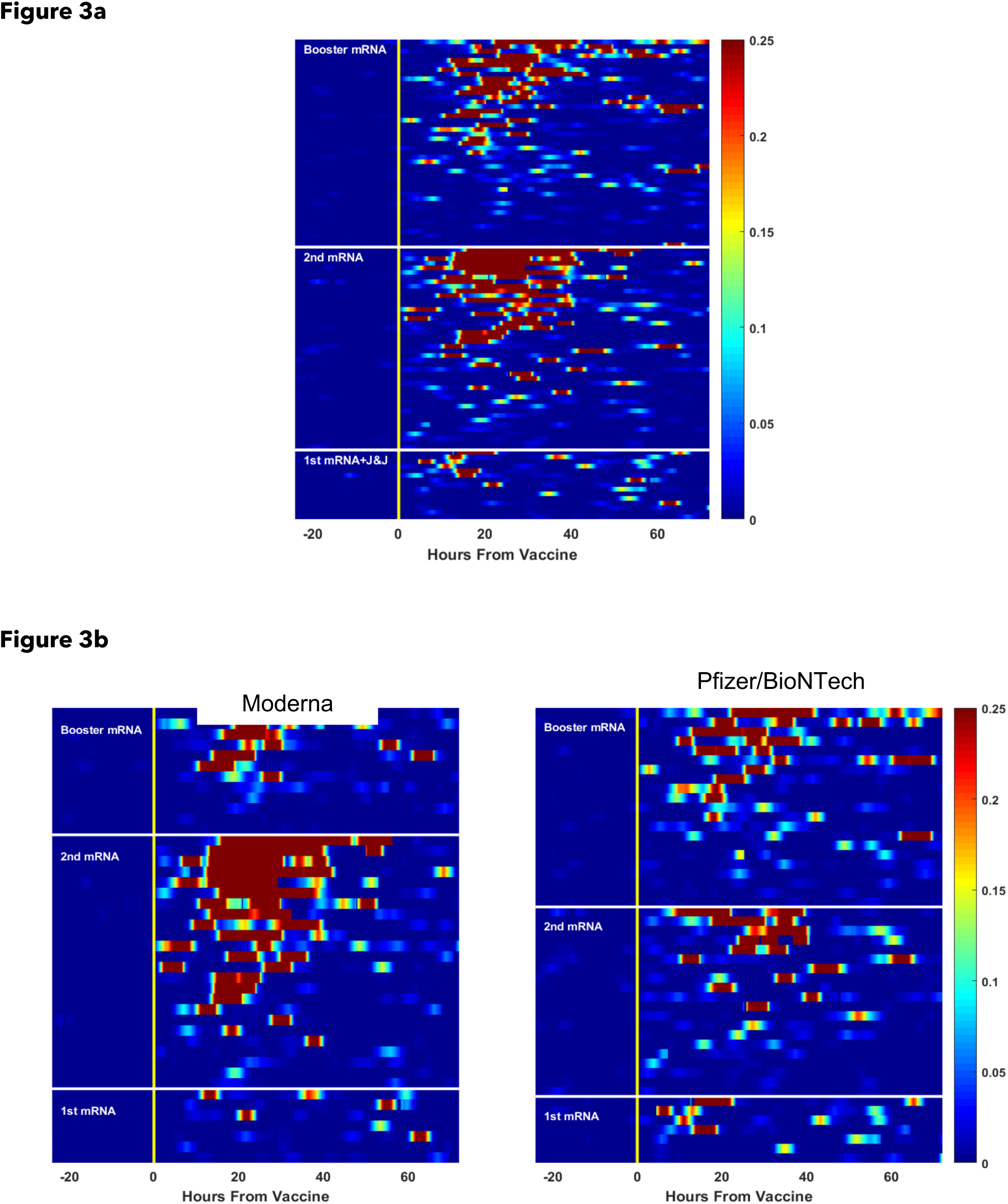
**a)** Heatmap showing inter-individual variation in the onset, degree, and duration of the inflammatory multivariate change index (iMCI) following 104 vaccine doses, aggregated by first, second or third vaccine dose. **b)** Similar heatmaps following 91 vaccine doses, aggregated by first, second or third vaccine dose for 43 Moderna and 48 Pfizer/BioNTech vaccine doses.

**Table 1:** Total post-vaccine inflammatory response following 104 vaccines as determined by the Area Under the iMCI curve based on vaccine dose, vaccine type, sex and age.

|  | n (%) with a detectable* response | Median (IQR) Area Under the iMCI Curve | p-value** of comparison of Area Under the iMCI Curve |
| --- | --- | --- | --- |
| 1<sup>st</sup> Dose (n=15) | 8 (53.3) | 0.043 (0.04) | 1<sup>st</sup> vs 2<sup>nd</sup> dose: p=0.01<br>1<sup>st</sup> vs 3<sup>rd</sup> dose: p=0.06<br>2<sup>nd</sup> vs 3<sup>rd</sup> dose: p=0.41 |
| 2<sup>nd</sup> Dose (n=44) | 31 (70.5) | 0.083 (0.18) |  |
| 3<sup>rd</sup> Dose (n=45) | 27 (60.0) | 0.063 (0.13) |  |
| mRNA-1273 (n=36)<br>(2<sup>nd</sup> or higher dose) | 26 (72.2) | 0.116 (0.19) | p=0.06 |
| BNT162b2 (n=41)<br>(2<sup>nd</sup> or higher dose) | 24 (58.5) | 0.055 (0.096) |  |
| Females (n=41)<br>(2<sup>nd</sup> or higher dose) | 25 (61.0) | 0.070 (0.15) | p=0.60 |
| Males (n=48)<br>(2<sup>nd</sup> or higher dose) | 33 (68.8) | 0.065 (0.14) |  |
| Lowest Tertile Age<br>(range 19-28) (n=31)<br>(2<sup>nd</sup> or higher dose) | 21 (67.7) | 0.070 (0.13) | Lowest vs Middle Tertile: p=0.99<br>Middle vs Highest Tertile: p=0.13<br>Lowest vs Highest Tertile: p=0.08 |
| Middle Tertile Age<br>(range 28-45.8) (n=26)<br>(2<sup>nd</sup> or higher dose) | 20 (76.9) | 0.056 (0.12) |  |
| Highest Tertile Age<br>(range 45.8 - 69) (n=29)<br>(2<sup>nd</sup> or higher dose) | 16 (55.2) | 0.077 (0.20) |  |

\* Detectable was defined as the presence of at least 50% of 15-minute steps within a 6-hour sliding window had an iMCIs > 0.10.
\*\* p-values obtained using Kolmogorov-Smirnov (KS) test

### Relationship of iMCI to Subjective Reactogenicity

Participants were asked to voluntarily self-report any symptoms following vaccination via an in-app survey. No data was entered following 15 of the 104 vaccine doses and were excluded from the analysis. Reported symptoms were classified as either systemic or local.^4^ Of the 89 doses with post-vaccine symptom data entered, a lack of any symptoms was documented following 10 doses (11.2%), local symptoms only following 9 (10.1%) doses, and systemic symptoms following 70 (78.6%) of doses. Of the 70 doses with systemic symptoms, 50 also reported local symptoms. Compared to vaccine doses associated with no systemic symptoms, those that were had a statistically significantly greater total inflammatory response as measured by AUC iMCI (median [IQR] 0.043 [0.039] vs 0.078 [0.14], p=0.008). **(Figure 4)**

**Figure 4.**
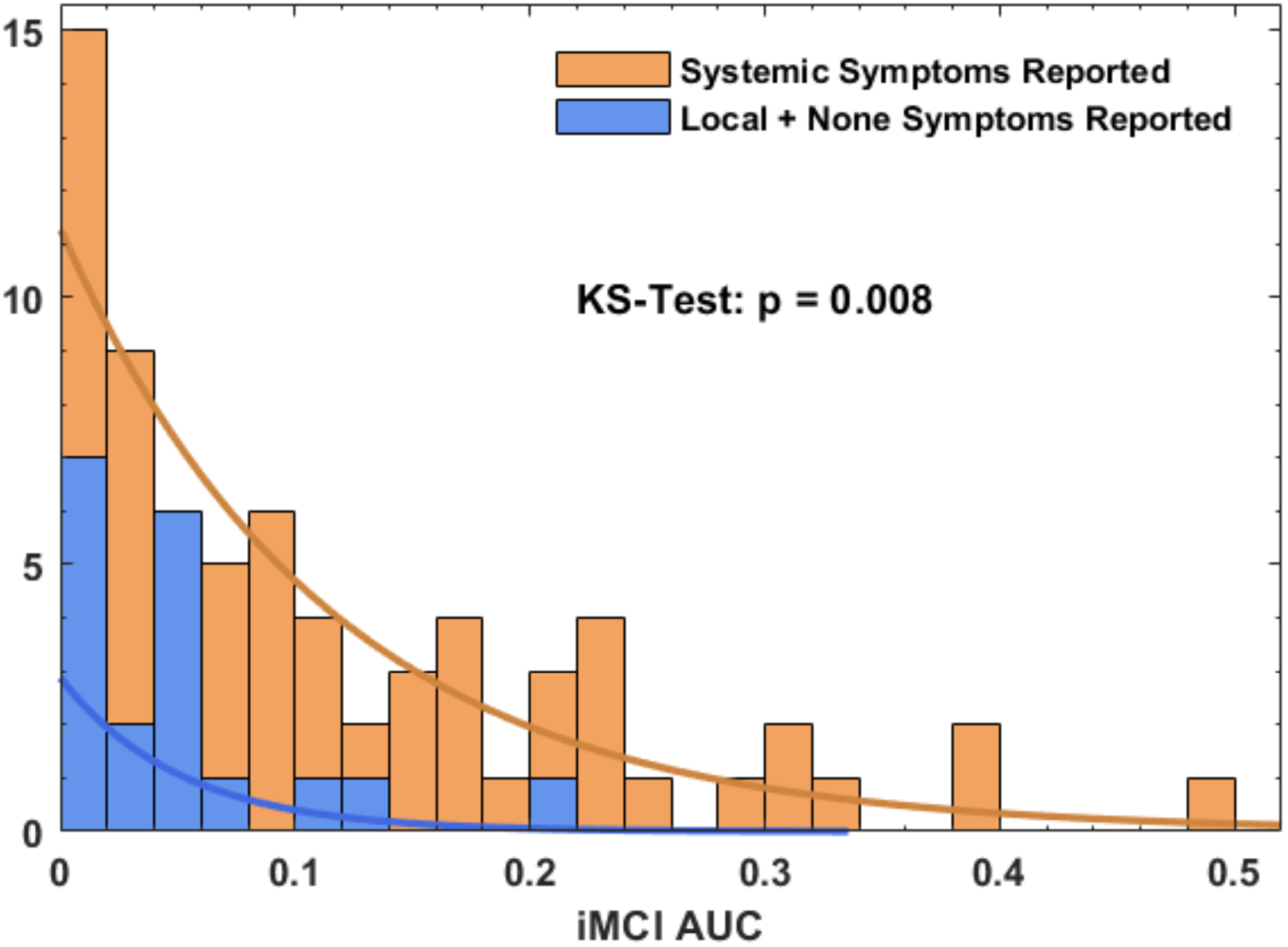
The distribution of AUC iMCI levels from 89 doses classified by whether they were associated with the participant experiencing systemic symptoms (70) or just local or no symptoms (19).

### Relationship of iMCI to Immunogenicity

Twenty-one individuals participated in the immunogenicity sub-study. Their mean age was 37.2 years, 65% were female, and 45% received the mRNA-1273 vaccine, and 55% BNT162b2. Twenty of the 21 sub-study participants had sufficient wearable data to determine their complete physiologic response following a second or booster vaccine dose. For these sub-study participants, the median AUC iMCI [IQR] was 0.055 [0.15], which was comparable to the overall population. Changes in both T-cell response and SARS-CoV-2 anti-spike protein IgG titers from baseline to day 14 were compared to the AUC iMCI response following vaccination.

The change in SARS-CoV-2 anti-spike protein IgG titer (ELU/mL) at day 14 following a 2^nd^ or 3^rd^ vaccine dose was significantly correlated with their AUC iMCI after the vaccine (Spearman *ρ*= 0.45, one-sided p=0.03) for the 19 individuals after exclusion of outliers. **(Figure 5a)** Similarly, the AUC iMCI was directly correlated with the increase in frequency of interleukin-21 expressing CD4+ cells (Spearman *ρ*= 0.56, one-sided p=0.009) at day 14 but was inversely correlated with the change in interferon-gamma expressing CD8+ cells (Spearman *ρ*= -0.47, one-sided p=0.029) for the 17 participants after exclusion of outliers. **(Figure 5b and c)**

**Figure 5.**
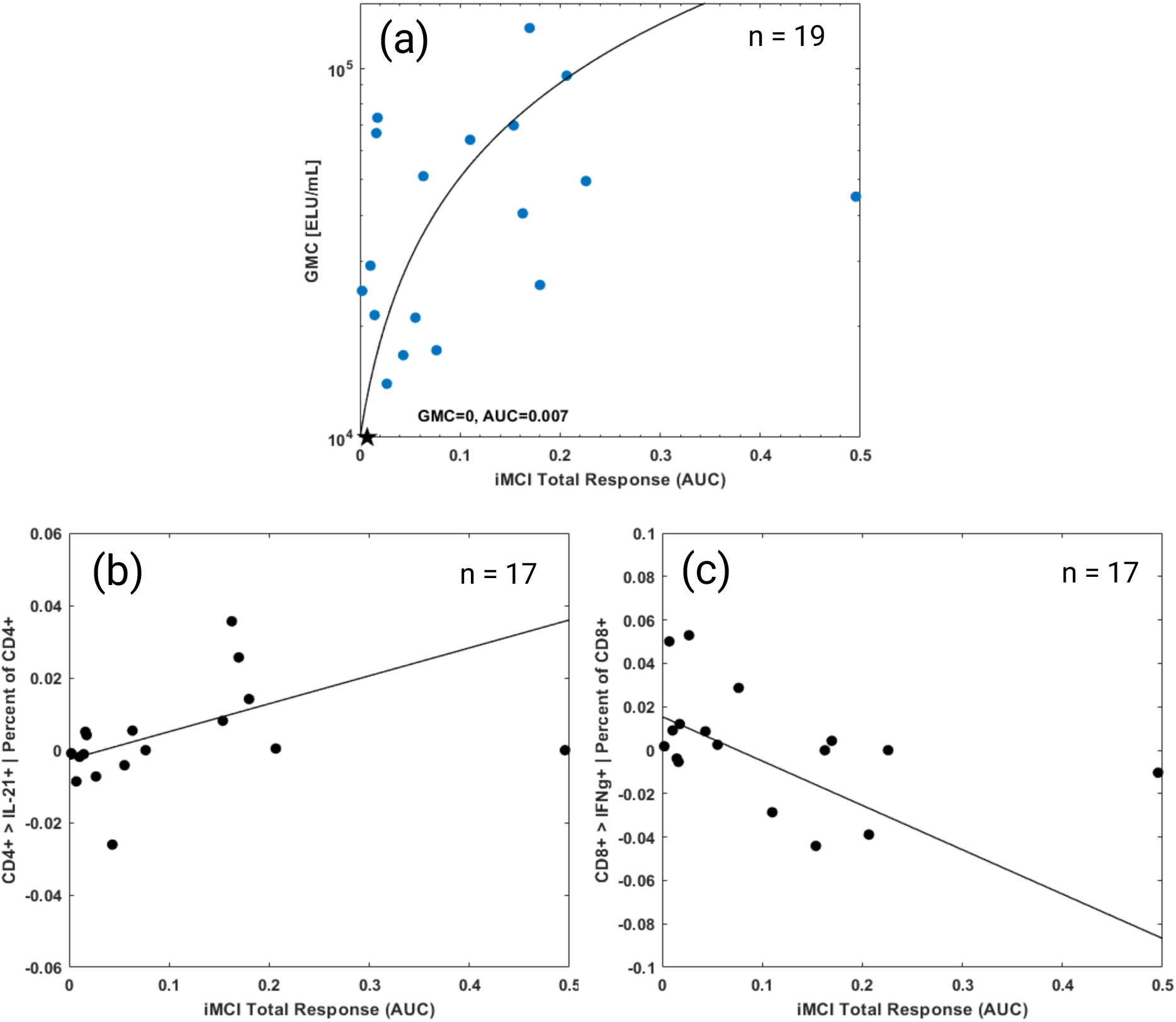
a) AUC iMCI total response versus change from baseline of Anti-SARS-CoV-2 anti-spike protein IgG titer in 19 participants following a 2^nd^ or 3^rd^ mRNA vaccine dose. The individual designated by the star had a lower titer at day 14 than at baseline. **b)** AUC iMCI total response versus change from baseline of frequency of interleukin-21 (IL-21+) expressing CD4+ cells and **c)** iMCI AUC total response versus change from baseline of frequency of interferon-gamma (IFN-γ+) expressing CD8+ cells in 17 participants14 days following 2^nd^ or 3^rd^ mRNA vaccine dose.

## Discussion

In this work, we characterized the interindividual heterogeneity in physiologic response to vaccination against COVID-19 collected via a medical-grade, wearable, continuous biosensor. Using these data and similarity-based modeling, we developed a digital biomarker, iMCI, that captures the entirety of an individual’s unique multivariate vaccine-induced physiologic response. As inflammatory response to a vaccine is currently only measurable at scale by tracking subjective symptoms of reactogenicity, an individualized inflammatory digital biomarker, once prospectively validated, could significantly enhance vaccine development and deployment.

Immune responses are known to vary greatly between people.^23^ Consistent with that, in the current study we identified substantial inter-individual variability in the physiologic response to vaccination that was, at least in part, consistent with known differences in vaccination-induced reactogenicity, such as less symptoms following a first does relative to subsequent doses, or greater symptoms in those receiving the mRNA-1273 versus BNT162b2 vaccine.^4^ We too found a significantly greater inflammatory response, as measured by AUC iMCI, following a second dose relative to a first dose. Those who received a mRNA-1273 vaccine as their second or third dose compared to those who received a BNT162b2 vaccine had a numerically greater inflammatory response, although not statistically significant. Similarly, while prior studies have found female versus male sex and younger versus older age to be associated with a greater reactogenicity,^24^ we were only able to show similar directionality for a sex difference, but not statistically significant differences in responses for sex or age, likely due to our small sample size.

In our small sub-study, we also found that an individual’s AUC iMCI response following vaccination was significantly associated with both humoral and cellular immune response at 14 days after vaccination. These results are consistent with multiple studies that have correlated higher degrees of vaccine-induced inflammation with a greater immune response. For example, using subjective reactogenicity as a measure of individual inflammatory response to vaccination, several studies have identified that more symptoms are associated with a greater humoral immune response.^25–28^ However, others have not.^29–31^ These inconsistent findings, and the known confounding by the nocebo affect,^5^ highlight the need for objective measures of individual inflammatory response to vaccination. To date, that has been limited to small studies measuring changes in serum cytokine levels following vaccination. Two such studies in individuals receiving the BNT162b2 vaccine found a correlation between the level of increase of multiple chemokines and cytokines following vaccination, especially interleukin-15 and interferon gamma, and subsequent Spike antibody levels.^7,32^

Multiple prior studies have also shown that subtle, individualized physiologic changes associated with vaccination against COVID-19 are detectable through wearable sensors. Most of these studies used consumer device sensor data, with biometrics typically summarized as a single data point per day. The majority used wrist- or ring-wearable data, evaluating changes post-vaccine in heart rate,^12,13,15–17^ respiratory rate,^13,15^ peripheral temperature,^13^ HRV,^13,15–17^ sleep,^12,15^ and activity.^12^ One additional study used a multiparametric sensor patch that in addition to the above parameters also included oxygen saturation, blood pressure, cardiac output and systemic vascular resistance.^14^ Our study adds to this prior work in several ways. First, we used a medical grade patch sensor that provided high-fidelity, beat-to-beat data that were processed to produce 17 source signals at a one-minute sampling rate. These data enabled a person’s unique physiologic changes associated with inflammation to be objectively tracked in 15-minute intervals. Our analytics incorporated all data streams and their interactions simultaneously using a machine learning method of similarity-based modeling to create a ‘digital twin’ of each participant.^33^ This allowed us to continuously compare monitored physiological signals with each participant’s baseline model of their unique dynamic physiologic patterns, which effectively removed expected activity-related, circadian and other personalized variations and left only vaccine-induced differences.

Just under 13.6 billion vaccine doses against COVID-19 have been administered globally through June of 2023.^34^ For the 67% of the world’s population who have received at least one dose of a vaccine, the overwhelming majority of them received the same dose, or series of doses, depending on the vaccine type, and unrelated to their individual underlying immune state. This is despite the fact that a host of personal characteristics influence the immune response to vaccination including age and sex,^35^ race,^36^ genetic and epigenetic factors,^37,38^ gut microbiome,^39^ sleep before, and time of day of vaccination,^40^ previous immune system exposures,^41^ and much more known and unknown.^42^ The inability to accurately predict a person’s response to vaccination is a major unmet need. Recent multi-omics and high-level transcriptional profiling studies in hundreds of individuals before and after vaccination have confirmed the complexity and broad heterogeneity in immune response to vaccination.^38,43^ In a study including data from 820 adults receiving one of 13 different vaccines, nearly two-thirds of the transcriptome variance was unexplained by identifiable clinical or vaccine characteristics.^44^

These results suggest that until a test is available that can reliably identify what an individual’s response to a vaccine will be prior to vaccination to enable proactive personalized dosing, a biomarker of what that individual’s response was to a received vaccine is needed.

Recognizing that a person didn’t experience the inflammatory response expected after a vaccine could potentially influence the timing or frequency of a booster dose. This might be especially important as personalized cancer vaccines continue to be developed.^45^ Following further validation, the personalized digital biomarker described here could enable a method to track an individual’s inflammatory response to vaccination using wearables. This could enable better objective tracking of reactogenicity and potentially serve as a surrogate for immunogenicity.

## Limitations

While there are advantages to the real-world study design in terms of eventual implementation, there are also multiple limitations. The largest limitation, especially for the development of an inflammatory biomarker, is the known marked heterogeneity in inflammatory response to vaccination and the lack of serum biomarkers to confirm that the physiologic changes detected post-vaccination are due solely to inflammation. Another limitation is that our analysis is limited to a patch sensor data. While the ECG provides higher quality heart rate data and its derivatives than do wrist- or ring-based photoplethysmography-based sensors, we are unable to determine if the greater data availability improves the clinical value in terms of quantifying reactogenicity or predicting immunogenicity. Finally, the inverse relationship between iMCI-detected inflammation and interferon-gamma expressing CD8+ cells is difficult to explain physiologically, which might suggest it is a chance finding.

## Conclusion

There is an unmet need to for a noninvasive method to monitor the totality of inflammation following vaccination to aid in the development and potential personalization of vaccines to enhance both safety and efficacy. In this work we show the potential for developing a personalized digital biomarker for vaccine-induced inflammation by combining medical-grade wearable sensor data and machine learning-enabled digital twin technology in the setting of real-world vaccination against COVID-19. We found that that our digital biomarker, iMCI, correlated with both subjective reactogenicity and immunogenicity. If confirmed in further studies, a personalized digital biomarker for inflammation could play an important role in improving vaccine safety and efficacy.

## Data Availability

The data obtained in the study is the property of the sponsor. Data sharing can be determined on a per case basis via request to authors.

## Conflicts of Interest

The current study was sponsored by physIQ, Inc. (Chicago, IL, USA) a company that has since been purchased by Prolaio, Inc. (Scottsdale, AZ, USA). A patent was filed by physIQ for the digital biomarker described. Some co-authors are employees of the company (JS, MG, SW) or a paid consultant (SRS). HG and EP are employees of CellCarta Bioscience, Inc. (Montreal, QC, Canada).

## Supplementary Information

**Supplemental Figure A:**
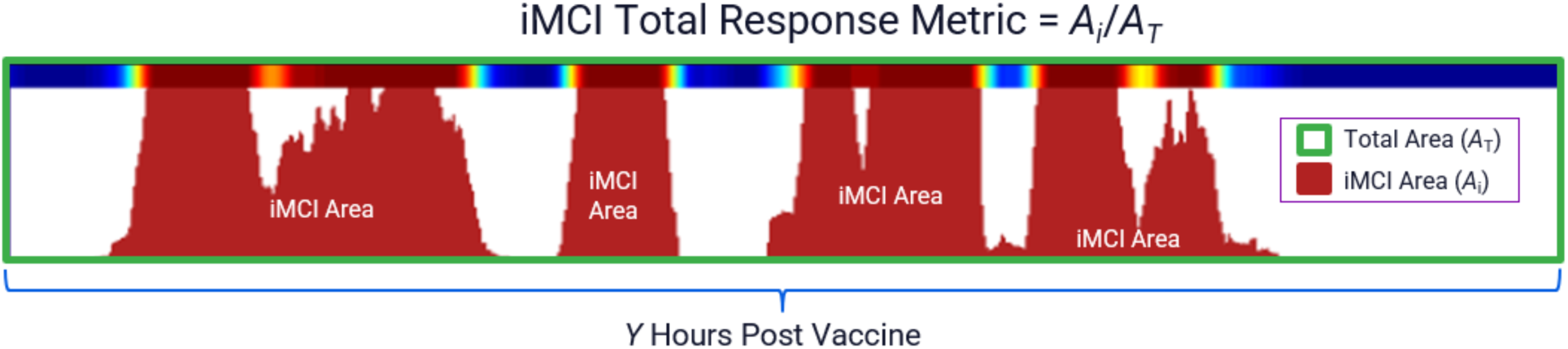
The iMCI Total Response was defined as A_i_/A_T_ as illustrated in the figure, where A_T_ is the total rectangular area within the time window (the green box) and A_i_ is the area under the curve for iMCI during the window (red area).

**Supplemental Figure B:**
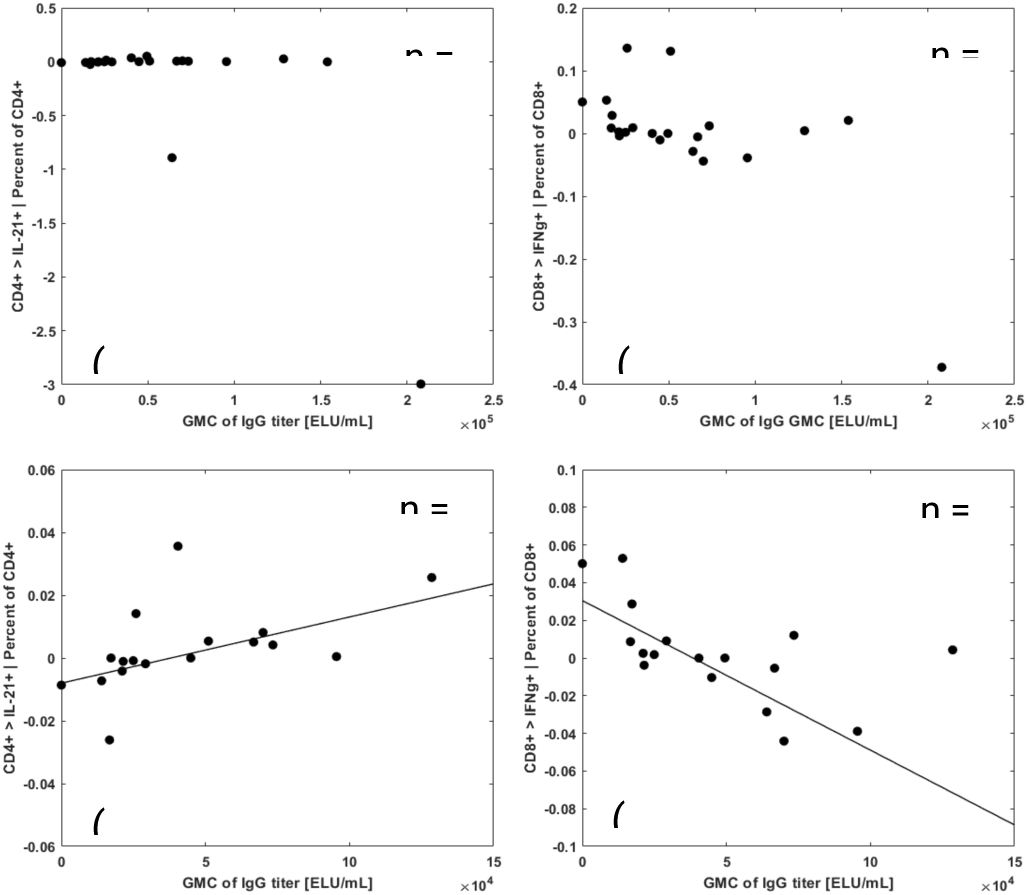
The two top figures (a) and (b) of Supplemental Figure (inserted above for reference) reveal an existence of few outliers. Using the statistical methods such as “median” for anti-spike protein IgG and “generalized extreme studentized deviate test” for T-cell assays we identified outliers and removed them from correlation analyses. There were two outliers in anti-spike protein IgG and three in case of each T-cell assay. One of outliers is common for both T-cell assays and anti-spike protein IgG. This resulted in 17 clinical data points for anti-spike protein IgG and T-cell assays to compare. The resulting correlation between anti-spike protein IgG and T-cell assays in clinical data after outlier removal is shown in two bottom figures (c) and (d) of Supplemental Figure (anti-spike protein IgG and T-cell assay IL-21+: 0.75, one-sided p=0.0003; anti-spike protein IgG and T-cell assay: -0.60, one-sided p=0.007).

## Notes

### Funding Statement

The study was funded by physIQ, Inc. (Chicago, IL, USA) a company that has since been purchased by Prolaio, Inc. (Scottsdale, AZ, USA). Some co-authors are employees of the company (JS, MG, SW) or a paid consultant (SRS).

### Author Declarations

The manuscript contains data from two near-identical prospective studies. The Sterling IRB gave ethical approval for the VIII study. The IRB of Purdue University gave ethical approval for the COmMON SENS study.

